# Serology-informed estimates of SARS-COV-2 infection fatality risk in Geneva, Switzerland

**DOI:** 10.1101/2020.06.10.20127423

**Authors:** Javier Perez-Saez, Stephen A Lauer, Laurent Kaiser, Simon Regard, Elisabeth Delaporte, Idris Guessous, Silvia Stringhini, Andrew S Azman, for the Serocov-POP Study Group

## Abstract

The infection fatality risk (IFR) is the average number of deaths per infection by a pathogen and is key to characterizing the severity of infection across the population and for specific demographic groups. To date, there are few empirical estimates of IFR published due to challenges in measuring infection rates.^1,2^ Outside of closed, closely surveilled populations where infection rates can be monitored through viral surveillance, we must rely on indirect measures of infection, like specific antibodies. Representative seroprevalence studies provide an important avenue for estimating the number of infections in a community, and when combined with death counts can lead to robust estimates of the IFR.

---

We estimated overall and age-specific IFR for the canton of Geneva, Switzerland using age-stratified daily case and death incidence reports combined with five weekly population-based seroprevalence estimates.^3^ From February 24th to June 2nd there were 5’039 confirmed cases and 286 reported deaths within Geneva (population of 506’765). We inferred age-stratified (5-9, 10-19, 20-49, 50-65 and 65+) IFRs by linking the observed number of deaths to the estimated number of infected individuals from each serosurvey. We account for the delays between infection and seroconversion as well as between infection and death.^4^ Inference is drawn in a Bayesian framework that incorporates uncertainty in seroprevalence estimates (supplement).

Of the 286 reported deaths caused by SARS-CoV-2, the youngest person to die was 31 years old. Infected individuals younger than 50 years experienced statistically similar IFRs (range 0.00032-0.0016%), which increases to 0.14% (95% CrI 0.096-0.19) for those 50-64 years old to 5.6% (95% CrI 4.3-7.4) for those 65 years and older (supplement). After accounting for demography and age-specific seroprevalence, we estimate a population-wide IFR of 0.64% (95% CrI 0.38-0.98).

Our results are subject to two notable limitations. Among the 65+ age group that died of COVID-19 within Geneva, 50% were reported among residents of assisted care facilities, where around 0.8% of the Geneva population resides. While the serosurvey protocol did not explicitly exclude these individuals, they are likely to have been under-represented. This would lead to an overestimation of the IFR in the 65+ age group if seroprevalence in this institutionalized population was higher than in the general population (supplement). Further, our IFR estimates are based on current evidence regarding post-infection antibody kinetics, which may differ between severe and mild infections. If mild infections have significantly lower and short-lived antibody responses, our estimates of IFR may be biased upwards.^5^

Estimates of IFR are key for understanding the true pandemic burden and for weighing different risk reduction strategies. The IFR is not solely determined by host and pathogen biology, but also by the capacity of health systems to treat severe cases. Despite having among the highest per capita incidence in Switzerland, Geneva’s health system accommodated the influx of cases needing intensive care (peak of 80/110 ICU-beds including surge capacity) while maintaining care quality standards. As such, our IFR estimates can be seen as a best-case scenario with respect to health system capacity. Our results reveal that population-wide estimates of IFR mask great heterogeneity by age and point towards the importance of age-targeted interventions to reduce exposures among those at highest risk of death.

## Data Availability

Data and code used in the analysis is available on Github

https://github.com/HopkinsIDD/sarscov2-ifr-gva

**^SEROCoV-POP STUDY Group:** Silvia Stringhini^1, 2, 3^, Idris Guessous^1, 2^, Andrew S. Azman^4,5^, Hélène Baysson^2^, Prune Collombet^1,2^, David De Ridder^2^, Paola d’Ippolito^1^, Yaron Dibner^1^, Natalie Francioli^1^, Kailing Marcus^1^, Chantal Martinez^1^, Natacha Noel^1^, Francesco Pennacchio^1^, Dusan Petrovic^1,3^, Attilio Picazio^1^, Giovanni Piumatti^1,8^, Jane Portier^1^, Caroline Pugin^1^, Barinjaka Rakotomiaramanana^1^, Aude Richard ^1,4^, Stephanie Schrempft^1^, Maria-Eugenia Zaballa^1^, Ania Wisniak^4^, Antoine Flahault^1,2,4^, Isabelle Arm Vernez^9^, Olivia Keiser^4^, Loan Mattera^17^, Magdalena Schellongova^2^, Laurent Kaiser^2,6,9,14^, Isabella Eckerle ^2,6,9^, Pierre Lescuyer^6^, Benjamin Meyer^2, 13^, Géraldine Poulain^6^, Nicolas Vuilleumier^2,6^, Sabine Yerly^6,9^, Sultan Bahta^2^, Jonathan Barbolini^2^, Rebecca Butzberger^2^, Sophie Cattani^2^, Alioucha Davidovic^2^, Antoine Daeniker^2^, Eugénie de Weck^18^, Céline Dubas^2^, Joséphine Duc^2^, Céline Eelbode^2^, Benoit Favre^2^, Alice Gilson^2^, Julie Guérin^2^, Lina Hassar^2^, Aurélia Hepner^2^, Francesca Hovagemyan^2^, Melis Kir^2^, Fanny Lombard^2^, Amélie Mach^2^, Eva Marchetti^2^, Soraya Maret^2^, Kourosh Massiha^2^, Virginie Mathey-Doret^2^, Tom Membrez^2^, Natacha Michel^2^, Emmanuelle Mohbat^2^, Hugo-Ken Oulevey^2^, Irine Sakvarelidze^2^, Milena Stimec^2^, Natacha Vincent^2^, Kor-Gaël Toruslu^2^, Nawel Tounsi^2^, Vincent^2^, Manon Will^2^, Alenka Zeballos Valle^2^, François Chappuis^1,2^, Delphine Courvoisier^1^, Laurent Gétaz^1,2^, Mayssam Nehme^1^, Febronio Pardo^22^, Guillemette Violot^23^, Sylvie Welker^1^, Alison Chiovini^1^, Odile Desvachez^16^, Benjamin Emery^2^, Acem Gonul^1^, Samia Hurst^7^, Gaëlle Lamour^21^, Yasmina Malim^1^, Philippe Matute^1^, Jean-Michel Maugey^22^, Aleksandra Mitrovic^1^, Didier Pittet ^12^, Klara M. Posfay-Barbe^2,10^, Jean-François Pradeau^22^, Christiane Rocchia Fine^1^, Lilas Salzmann-Bellard^1^, Mélanie Seixas Miranda^15^, Michel Tacchino^22^,Carol Theurillat^1^, Sophie Theurillat^21^, Mélissa Tomasini^1^, Didier Trono^11^, Zoé Waldman^2^

1. Division of Primary Care, Geneva University Hospitals, Geneva, Switzerland
2. Faculty of Medicine, University of Geneva, Geneva, Switzerland
3. University Centre for General Medicine and Public Health, University of Lausanne, Lausanne, Switzerland
4. Institute of Global Health, Faculty of Medicine, University of Geneva, Geneva, Switzerland
5. Department of Epidemiology, Johns Hopkins Bloomberg School of Public Health, Baltimore, USA
6. Division of Laboratory Medicine, Geneva University Hospitals, Geneva, Switzerland
7. Institut Ethique, Histoire, Humanités, University of Geneva, Geneva, Switzerland
8. Faculty of BioMedicine, Università della Svizzera italiana, Lugano, Switzerland
9. Geneva Center for Emerging Viral Diseases and Laboratory of Virology, Geneva University Hospitals, Geneva, Switzerland
10. Division of General Pediatrics, Geneva University Hospitals, Geneva, Switzerland
11. School of Life Sciences, Ecole Polytechnique Fédérale de Lausanne (EPFL), Lausanne, Switzerland
12. Infection Prevention and Control program and World Health Organization (WHO) Collaborating Centre on Patient Safety, Geneva University Hospitals, Geneva, Switzerland
13. Centre for Vaccinology, Department of Pathology and Immunology, University of Geneva, Geneva, Switzerland
14. Division of Infectious Diseases, Geneva University Hospitals, Geneva, Switzerland
15. Division of Diagnostics, Geneva University Hospitals, Geneva, Switzerland
16. Division of Women, Children and Adolescents, Geneva University Hospitals, Geneva, Switzerland
17. Campus Biotech, Geneva, Switzerland
18. Education Structure, University of Geneva, Geneva, Switzerland
19. Institute of Social and Preventive Medicine, Bern, Switzerland
20. Deutsches Primatenzentrum (DPZ), Göttingen University, Göttingen, Germany
21. Human Ressources Departement, Geneva University Hospitals, Geneva, Switzerland
22. Information Systems Division, Geneva University Hospitals, Geneva, Switzerland
23. Division of Communication, Geneva University Hospitals, Geneva, Switzerland

